# A Reproducible Protocol to Assess Arrhythmia Vulnerability in Silico: Pacing at the End of the Effective Refractory Period

**DOI:** 10.1101/2021.01.21.21250205

**Authors:** Luca Azzolin, Steffen Schuler, Axel Loewe, Olaf Dössel

## Abstract

In both clinical and computational studies, different pacing protocols are used to induce arrhythmia and non-inducibility is often considered as the endpoint of treatment. The need for a standardized methodology is urgent since the choice of the protocol used to induce arrhythmia could lead to contrasting results, e.g., in assessing atrial fibrillation (AF) vulnerabilty. Therefore, we propose a novel method – pacing at the end of the effective refractory period (PEERP) – and compare it to state-of-the-art protocols such as phase singularity distribution (PSD) and rapid pacing (RP) in a computational study. All methods were tested by pacing from 227 evenly distributed endocardial points in a bi-atrial geometry. 6 different atrial models were implemented: 4 cases without specific AF-induced remodelling but with decreasing global conduction velocity and 2 persistent AF cases with an increasing amount of fibrosis resembling different substrate remodeling stages. Compared with PSD and RP, PEERP induced a larger variety of arrhythmia complexity requiring, on average, only 2.7 extra-stimuli and 3 s of simulation time to initiate reentry. Moreover, PEERP and PSD were the protocols which unveiled a larger number of areas vulnerable to sustain stable long living reentries compared to RP. Finally, PEERP can foster standardization and reproducibility, since, in contrast to the other protocols, it is a parameter-free method. Furthermore, we discuss its clinical applicability. We conclude that the choice of the inducing protocol has an influence on both initiation and maintenance of AF and we propose and provide PEERP as a reproducible method to assess arrhythmia vulnerability.

## 1 Introduction

Atrial fibrillation (AF) is the most frequent cardiac arrhythmia and a progressive pathology associated with high morbidity and mortality [14]. Despite recent advances in both diagnostic and therapeutic techniques, the success rate for the standard-of-care treatment, catheter ablation, is sub-optimal in patients with persistent AF [33], [40]. The modest efficacy reflects the complexity of the underlying phenomena and our incomplete understanding of the mechanisms of initiation, maintenance and progression of AF episodes [2], [29]. In clinical practice, electrical stimulation has been widely used to diagnose and guide therapy of arrhythmias. Despite the high prevalence of atrial rhythm disorders, the sensitivity, specificity and reproducibility of mostly ventricular stimulation has been investigated [10], [21]. An equally critical evaluation of protocols for induction of AF and atrial flutter is currently lacking.

[22] highlighted the importance of the choice of the protocol used to induce AF even in patients without AF history and/or structural heart disease. The incidence of initiation and maintenance of AF varied according to gender, method of induction and number of inductions. In addition, the stimulation sites, pacing methods, number of AF inductions, use of pharmacological provocation and the definition of inducibility based on AF duration vary among different studies [11], [12], [16], [36]. Currently, various protocols are used to test AF inducibility before and after ablation procedures. For instance, some groups have used burst pacing at a fixed cycle length, while others have paced at the shortest cycle length which resulted in loss of 1:1 capture. [22] observed that the incidence of inducible or sustained AF was significantly higher with decremental pacing compared to burst pacing, as was the total duration of induced AF. They concluded that the adoption of AF inducibility as final electrophysiological endpoint is critically dependent on the variations in the definition of inducibility, aggressiveness of AF induction protocol and the number of AF inductions. However, they could not compare burst versus decremental pacing within the same patient as this is not feasible clinically for ethical reasons. In another clinical study [17], a programmed atrial stimulation protocol for induction of sustained arrhythmia was evaluated. They were able to induce and maintain arrhythmic episodes using a train of pulses close to the effective refractory period in 39/44 (89%) patients. The study demonstrated that employing only two atrial sites and three atrial extra-stimuli induced either AF or atrial flutter in 89% of the patients with previous AF history and in 7% of the control group without documented arrhythmias. Moreover, inducibility was proposed as a predictor of long-term AF recurrence [11].

Computational modelling has been proven to be a useful tool for assessing arrhythmia vulnerability [3], [4], [44] and for supporting ablation planning [6], [24], [28], [38]. However, different protocols used to induce arrhythmia in simulations [6], [20], [30], [37], [44] are not only making studies difficult to compare, but are also influencing the decision on whether an atrial model is vulnerable to AF and are therefore crucial for identifying the optimal ablation targets. In [6], arrhythmic episodes were induced by pacing from 40 evenly distributed sites applying a train of 12 electrical stimuli with decreasing cycle length from 300 to 150 ms. Their choice of protocol was motivated by a previous publication [37], in which, however, AF was initiated by delivering 5 triggering ectopic beats from the right superior pulmonary vein with a fixed coupling interval of 400 ms with the sinus rhythm but variable basic cycle length of 155 or 160 ms, depending on inducibility. Conversely, [30], proposed to induce AF by manually placing 1-6 phase singularities on the atrial surface, reconstructing an activation time map using an interpolation algorithm based on the eikonal equation and using that as initial condition for a monodomain simulation.

Considering the increasing use of catheter ablation and pacing therapies for atrial arrhythmias, which often take the suppression of inducibility as one of the endpoints of treatment, the definition of a commonly acknowledged electrical stimulation protocol is needed. A standardized methodology is critical since the use of different protocols could lead to contrasting conclusions on whether or not a patient or a patient-specific digital twin model is vulnerable to arrhythmia. In this study, we quantitatively evaluate different methods to induce arrhythmia, investigate their impact on both initiation and maintenance of AF episodes and propose an easily reproducible protocol to assess AF vulnerability of a particular atrial model.

## 2 Materials and Methods

### 2.1 Atrial Models

In this work, a highly detailed volumetric bi-atrial geometry derived from magnetic resonance images was used [18]. The tetrahedral mesh had an average edge length of 0.5 mm. Fiber orientation was calculated by a semi-automatic rule-based algorithm [43]. Different conductivity and anisotropy values were implemented in working myocardium, pectinate muscles, bachmann bundle, inferior isthmus and crista terminalis to consider the heterogeneity in the atria [25], [26]. Conductivities were tuned to reach 4 different total activation times in sinus rhythm of 130, 151, 179 and 199 ms and respective global average conduction velocities of 0.7, 0.66, 0.55 and 0.49 m/s, respectively. We will refer to these models as H1, H2, H3 and H4. The total activation times were chosen to be all ≥ 130 ms since patients with a P-wave duration longer than 130 ms were shown to have higher risk for AF [23], [34]. Depending on the simulated scenario, the original [9] model (H1, H2, H3 and H4) or a variant reflecting AF-induced remodeling (UII and UIV) [27] represented the myocyte membrane dynamics. Fibrotic tissue was included in the regions of both atria most-often exhibiting fibrotic substrate in patients with AF [1], [5], [7], [13]. Two different fibrosis severity stages were implemented: Fig. 1A shows the Utah stage II case (UII), in which 19% of the left atrial wall (LAW) and 5% of the right atrial wall (RAW) were modeled as fibrotic. In the most severe case (Fig. 1B), 39% of the LAW and 11% of the RAW was modelled as fibrotic, classifying as Utah stage IV (UIV). To account for structural remodeling and the presence of scar tissue, we set 50% of the elements in the fibrotic regions as almost not conductive (conductivity of 10^−7^ S/m). In the other 50%, several ionic conductances were rescaled to consider effects of cytokine-related remodeling [37] (−50% g_*K*1_, -40% g_*Na*_ and -50% g_*CaL*_).

**Figure 1:**
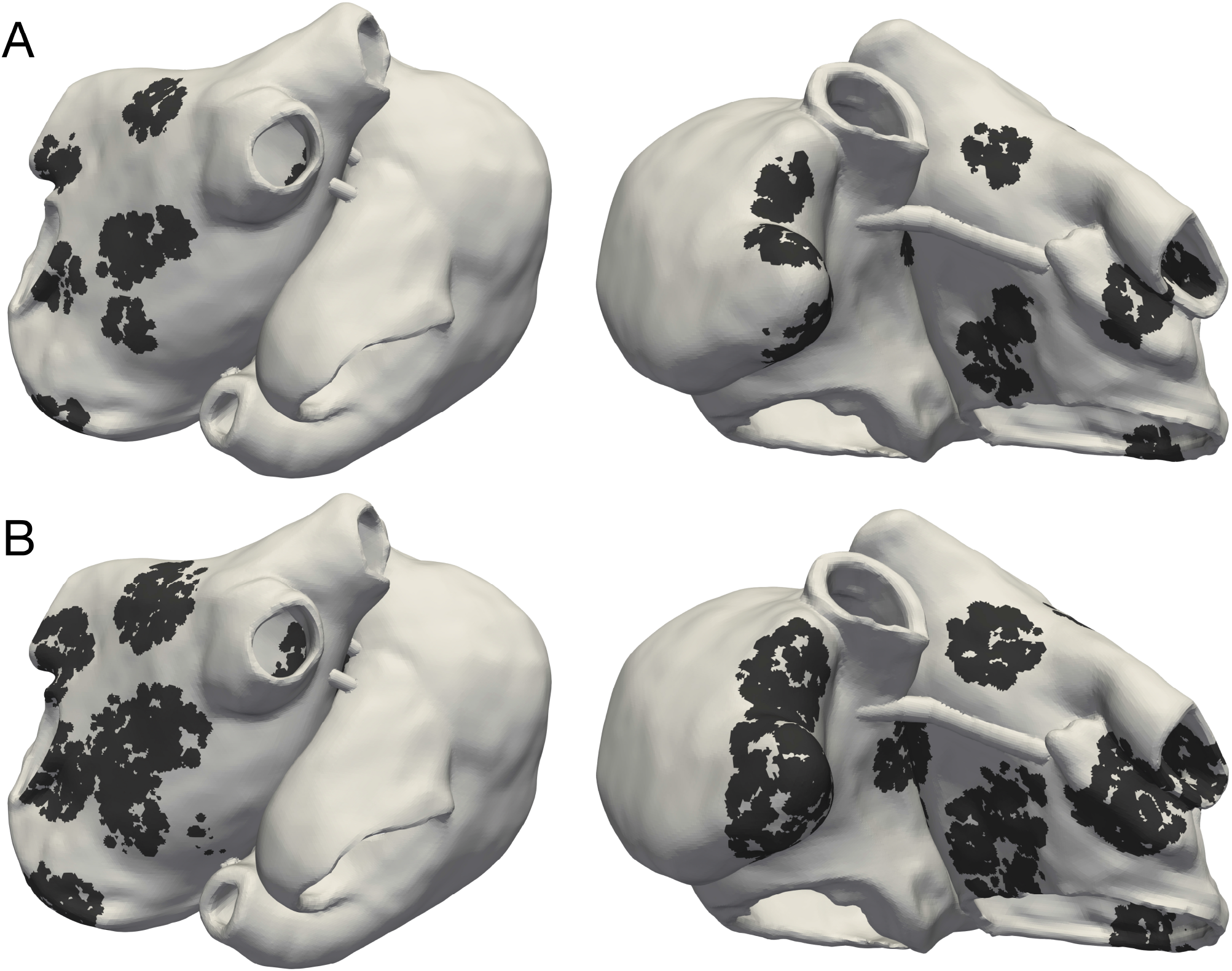
Posterior and anterior view of Utah stage II (UII) model in **(A)** and Utah stage IV (UIV) model in **(B)**. Black regions indicate fibrotic tissue in which 50% of the elements were non-conductive and the remaining 50% electrophysiologically remodeled due to cytokine effects.

### 2.2 Protocols to Test Inducibility

We tested inducibility of arrhythmic episodes by pacing from up to 227 evenly distributed points on the endocardial surface with an inter-point distance of 1 cm. We systematically evaluated and compared three state-of-the-art methods: phase singularity distribution (PSD), rapid pacing (RP) and pacing at the end of the effective refractory period (PEERP). We considered a point to be ‘inducing’ if the application of one protocol at that location, induced and sustained an arrhythmia for at least 1.5 s after the end of the protocol. All the protocols used in this study are available open source in the examples section of openCARP [39], [42] at www.opencarp.org/documentation/examples.

#### 2.2.1 Phase Singularity Distribution

The phase singularity distribution (PSD) method places phase singularites (PSs) in the atria, constructs an activation time map by solving the Eikonal equation and finally uses this as initial state for a monodomain excitation propagation simulation [15], [30]. The parameters of this method are the cycle length of the re-entrant wave, the number of PSs to initialize and the rotation direction of each PS. We set a single phase singularity rotating in anti-clockwise direction (looking from the endo-to the epicardium) in one of the 227 points for each of the 227 simulated scenarios. The cycle length was set to 315 and 168 ms, for the original [9] model and the chronic AF variant [27], respectively (5% longer than the effective refractory period).

#### 2.2.2 Rapid Pacing

The rapid pacing (RP) protocol consists of a train of stimulation pulses with decreasing coupling intervals (CI). We extended the protocol by supporting a variable number of pulses with the same CI before the next decrement. Moreover, we implemented the option to check for successful initiation of an arrhythmia after every stimulation instead of only at the end of the protocol. This extended RP protocol is defined by 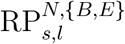 where *N* is the number of stimuli with the same CI, {*B, E*} defines if we were checking for the induction of an arrhythmia after every beat (*B*) or only at the end of the protocol (*E*), *s* is the starting CI and *l* is the last CI, both in ms. The CI was continuously decremented from *s* to *l* in steps of 10 ms. *s* was different for each action potential phenotype: 300 ms for the control case and 200 ms in the AF-induced remodelling setup. *l* was also distinct between control and AF remodelled and chosen as 200 ms and 130 ms, respectively. *N* was incremented from 1 to 4. For example, the RP protocol with *N* = 1, *s* = 200 ms, *l* = 130 ms and arrhythmia checking only at the end resulted in the following train of pulses:

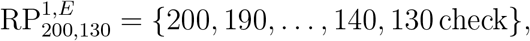

which is the most common RP protocol [6], [20], [44]. The case *N* = 2 was as follows:

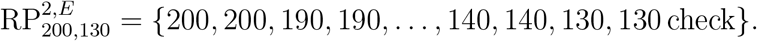

The RP protocol with *N* = 1, *s* = 200, ms *l* = 130 ms and arrhythmia checking after every beat yields:

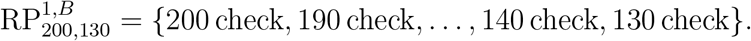

The parameters of the RP protocol are the maximum number of beats with the same CI, first and last CI, the CI decrement and the stopping criteria (arrhythmia checking in between or at the end of the protocol).

#### 2.2.3 Pacing at the End of the Effective Refractory Period

The pacing at the end of the effective refractory period (PEERP) protocol triggers stimuli at the end of the effective refractory period. Therefore, each stimulus was delivered as soon as the underlying tissue had recovered from the previous activation and was able to initiate a new wave propagation. We implemented a run-time binary search method to find the minimum time at which a new depolarization wave could locally spread (effective refractory period) with a temporal resolution of 1 ms. Based on initial experiments, the atrial action potential duration at 94% repolarization computed during sinus rhythm was chosen as first guess of the effective refractory period. The criterion for successful wave propagation was a transmembrane potential ≥ −50 mV in at least one node in a ring 4-6 mm around the stimulation site. In this study, the maximum number of beats delivered from one stimulation site was set to 4. Consequently, the stimulation at one pacing location was stopped if the maximum number of beats was reached or if an arrhythmia was initiated. The only parameter for this new PEERP protocol is therefore the maximum number of stimuli to apply at a specific location.

### 2.3 Arrhythmia Classification

Reentrant dynamics can, in many cases, be simplified by considering the behavior of their organising centers – the PSs. We followed the method presented by [8] to detect PSs and track the spatio-temporal behavior of each PS during the whole simulated episode. We defined a PS as long-living PS (llPS) if it was sustained for at least 500 ms. Moreover, we distinguished the llPS which were stable within an area enclosed in a bounding box of 5 cm edge length (stable llPS) for the last simulated 1.5 s, from the ones which were meandering largely (non-stable llPS). The episodes in which no llPS was found were classified in multiple wave fronts (Multi) or flutter (Fl) by visual inspection. We, therefore, classified each arrhythmic episode into 4 categories: Multi, Fl, non-stable llPS and stable llPS. The Multi class was defined as a complex and disorganised series of multiple wave fronts merging and colliding with each other without a clearly identifiable llPS. Periodic macro-reentries were interpreted as Fl cases. An example of stable llPSs and multiple wave fronts episodes are shown in Fig. 2.

**Figure 2:**
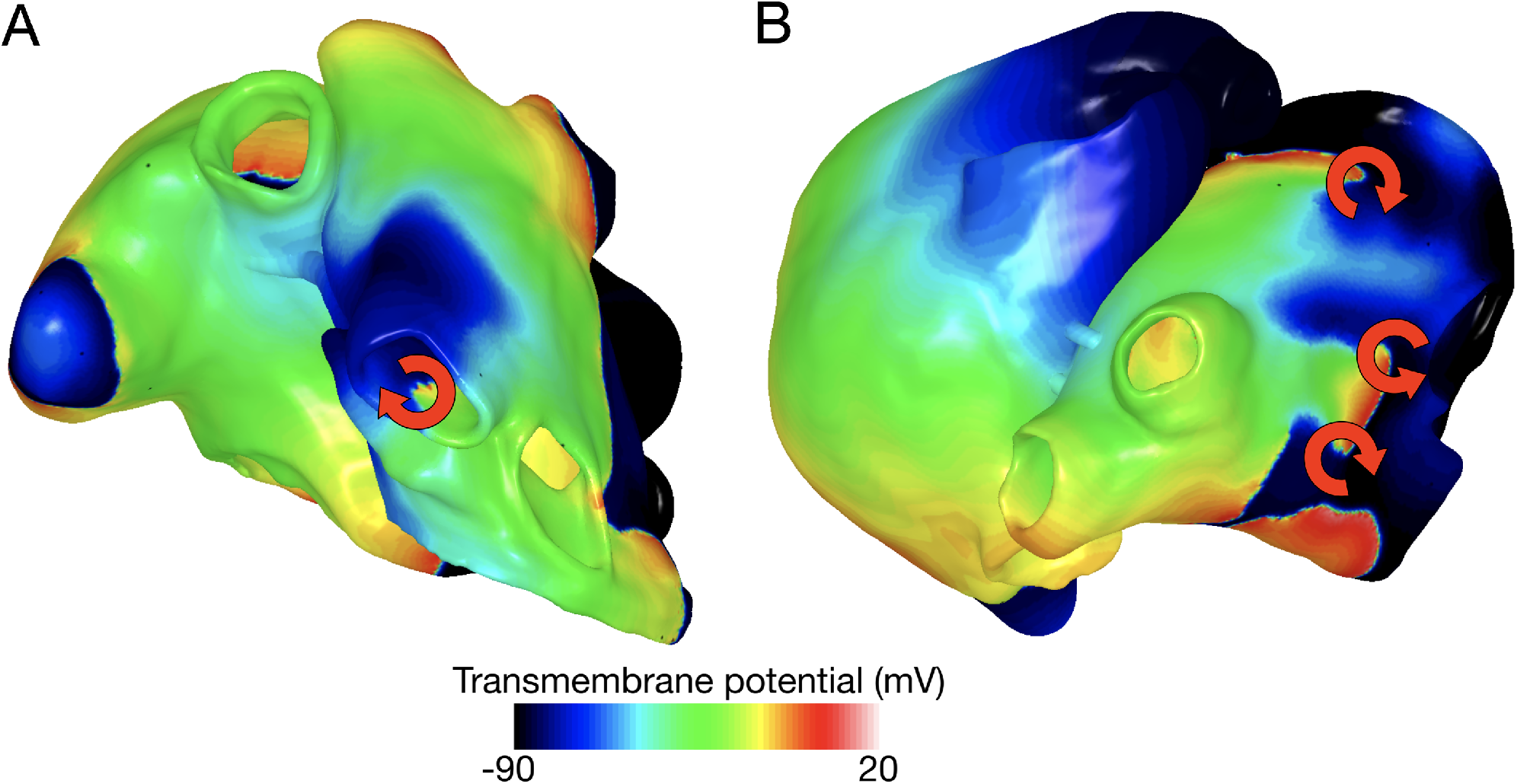
Example of stable llPS **(A)** and multiple wave fronts **(B)**.

### 2.4 Subdivision of Atria in Spatial Segments

To localize the inducing points and drivers sustaining atrial activity, we partitioned the atria into different regions. The atria were subdivided into 28 segments (Fig. 3): 19 in the left atrium (4 pulmonary vein segments (LIPV, RIPV, LSPV and RSPV), 4 segments at the roof, 5 segments on the posterior wall, 2 segments in the septum, 4 segments on the anterior wall) and 9 in the right atrium (inferior and superior vena cava rings, coronary sinus, cavotricuspid isthmus and septum, sinus node, right atrial appendage, anterior wall, 4 segments on the posterior wall of the RA).

**Figure 3:**
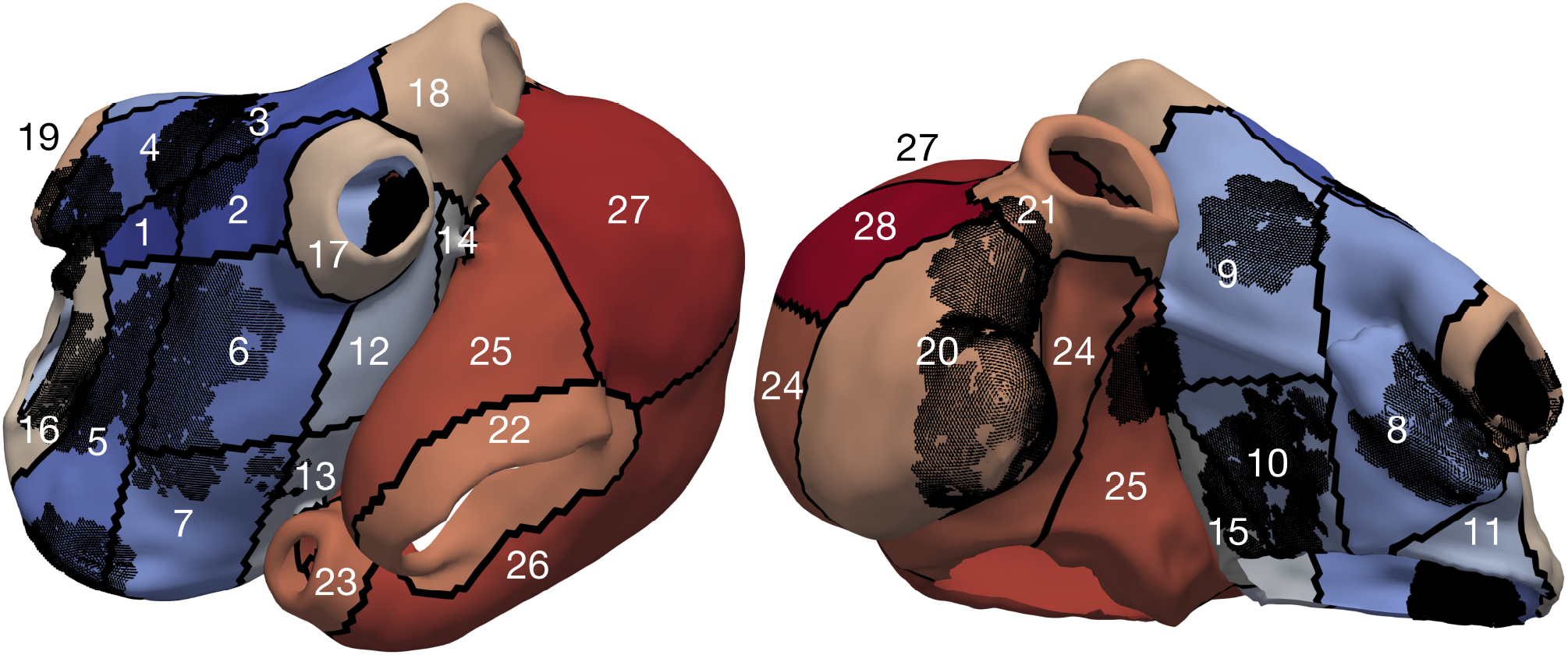
Model UIV partitioned into segments, the black dots represent the fibrotic elements.

### 2.5 Computational Tools

The spread of the electrical activation in the atrial myocardium was simulated by solving the monodomain system using openCARP [39], [42] and a time step of 0.02 ms.

## 3 Results

### 3.1 Inducibility

The number of inducing points and the relative frequency of arrhythmic mechanisms induced in each model are shown in Fig. 4. The number of inducing points increased with both a lower average global conduction velocity and higher amount of fibrotic tissue. No arrhythmic episode could be induced and sustained in model H1, which is why this model is not included in the results. PEERP was the only method which induced all the possible arrhythmic mechanisms described in Sec. 2.3 in most of the models (4/5). The application of the RP protocol resulted in inducing a majority of atrial flutter mechanisms in the model UII and no multi-frontal episode in both models with fibrosis. Moreover, the PEERP method initiated arrhythmia when pacing from the highest number of segments, averaging all over the models, as displayed in Fig. 5. However, RP^*B*^ was the only method which could induce and sustain arrhythmia pacing from all the segments in the UIV model. Nonetheless, inducing points belonging to only 4/28 segments were found applying the RP protocol in the model UII. The atrial segments in which inducing points were found to initiate and maintain stable llPSs applying the different protocol can be found in the supplementary material.

**Figure 4:**
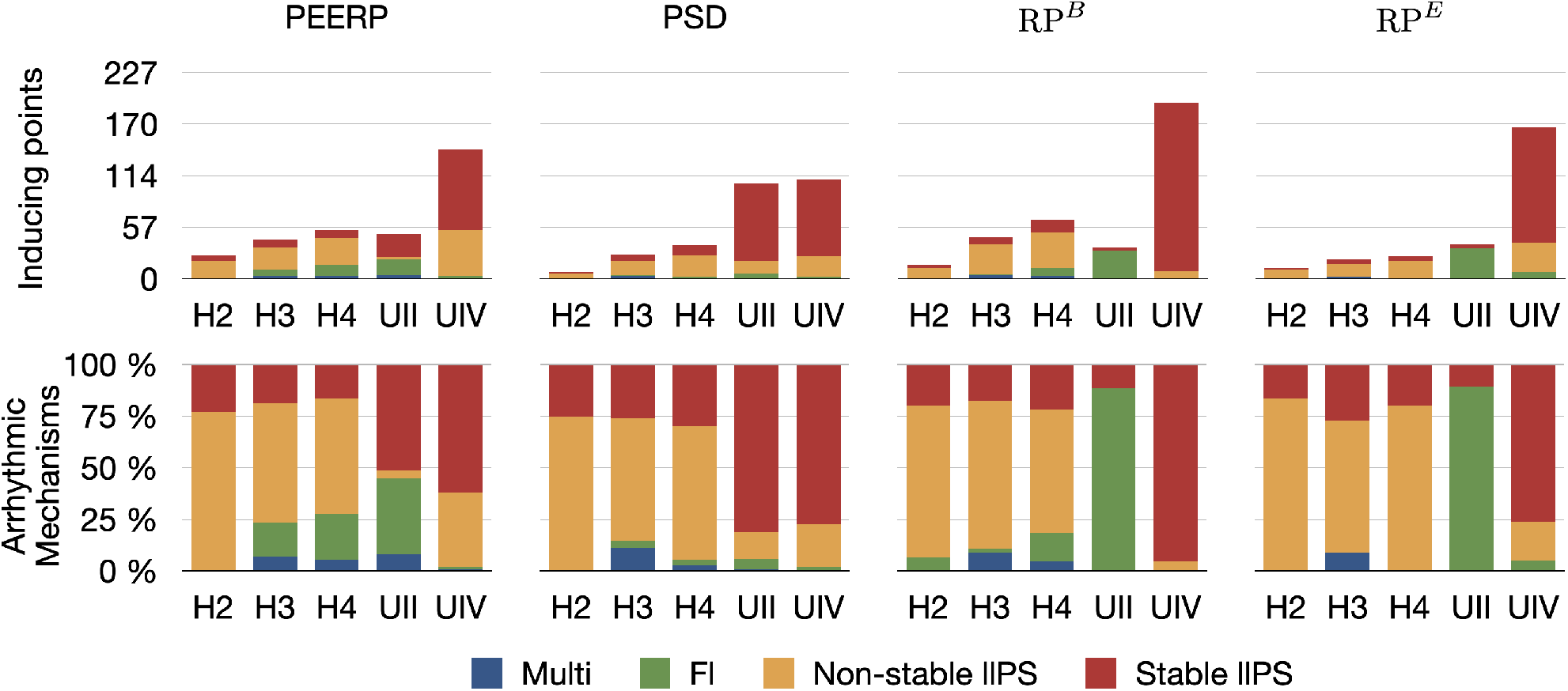
First row: Number of inducing points classified per induced arrhythmic mechanism: multiple wave fronts, flutter, non-stable and stable long living phase singularity. Second row: arrhythmic mechanisms induced by each protocol in the different atrial models.

**Figure 5:**
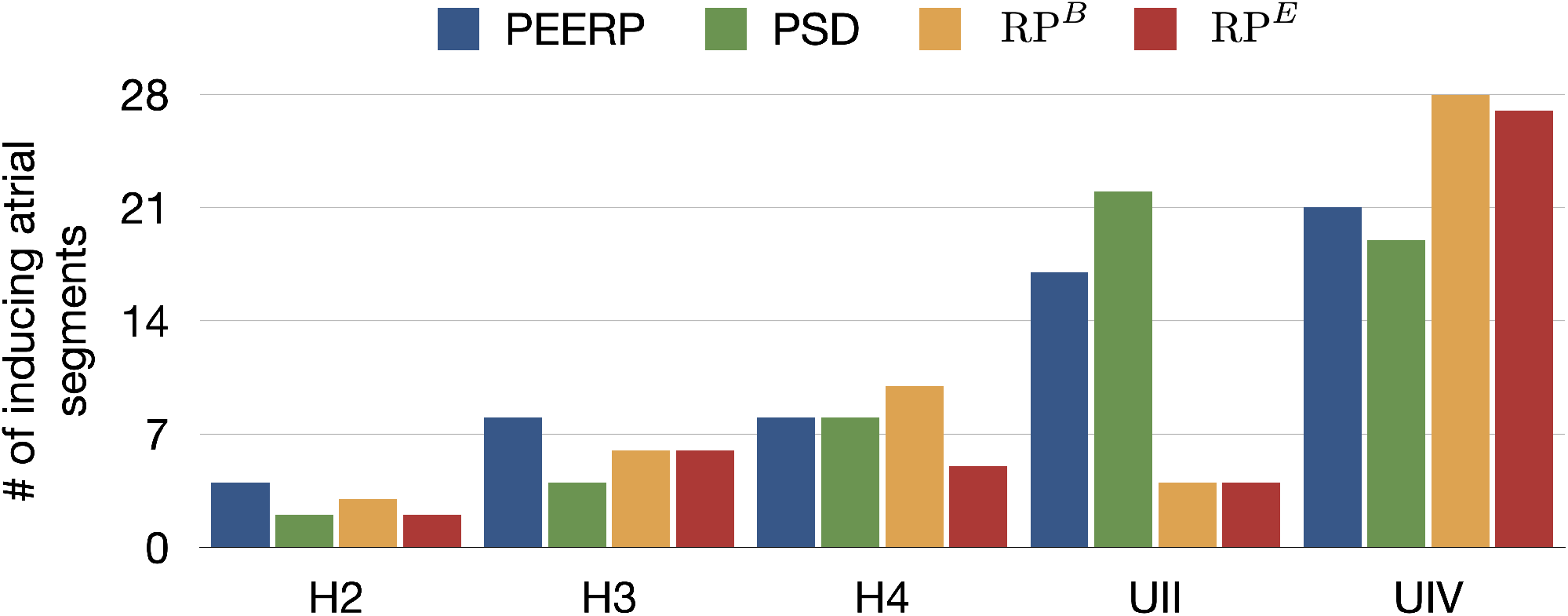
Number of atrial segments in which inducing points were identified applying the various protocols in the different models.

The CI of RP^*B*^ at which arrhythmia was initiated was around 130 ms and 250 ms for the models with and without structural remodelling, respectively (Fig. 6A). The following tendency was observed: a slight increase of the inducing CI with lower global conduction velocity and higher amount of fibrosis. The inducing number of stimuli with the same CI (*N*) was in most of the cases *N* = 2 for both RP^*B*^ and RP^*E*^, as presented in Fig. 6B. However, in the model UIV *N* = 1 was often sufficient to initiate arrhythmic episodes using RP^*B*^. The required total number of stimuli to induce arrhythmia by each protocol is displayed in Fig. 6C. The methods PEERP, RP^*B*^ and RP^*E*^ required on average 2.7, 13.6 and 17.6 beats respectively to initiate an arrhythmic episode.

**Figure 6:**
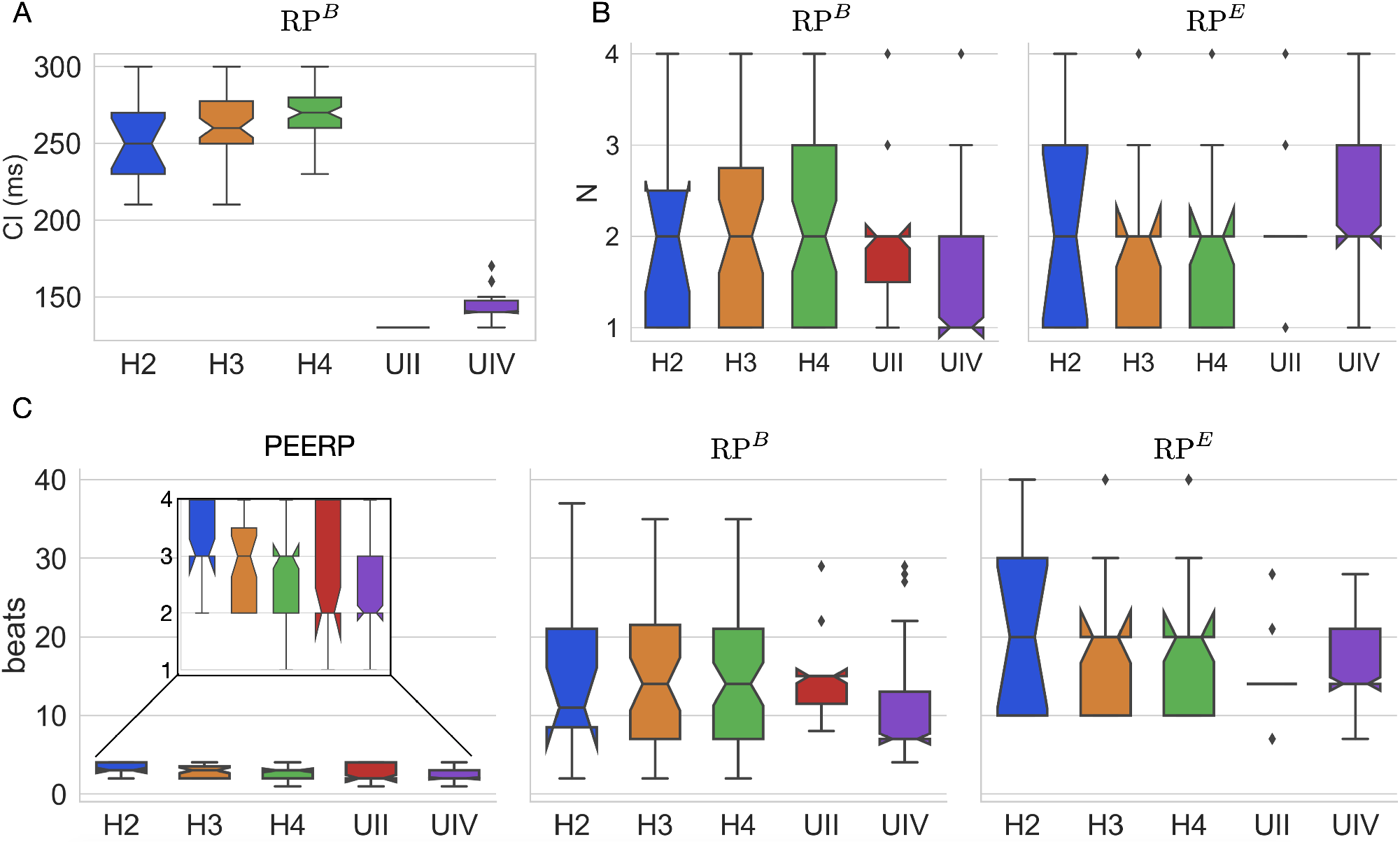
**(A)** CI at which RP^*B*^ induced arrhythmia in each atrial model. **(B)** number of beats per CI (*N*) needed to initiate arrhythmia in both RP^*B*^ and RP^*E*^. **(C)** total number of beats applied in the protocols PEERP, RP^*B*^ and RP^*E*^ inducing arrhythmic episodes.

Furthermore, the application of PEERP resulted in an increasing number of segments in which inducing points were found with both decreased global CV and greater amount of fibrotic tissue, as shown in Fig. 5. On one hand, the RP protocols were the only ones to initiate and sustain arrhythmia pacing from almost all the segments in the model UIV. On the other hand, both RP^*B*^ and RP^*E*^ could induce pacing from only 4 segments in the model UII.

Moreover, the simulation time required to complete each protocol for one stimulation point was 3.0 s, 19.5 s, 8.4 s for PEERP, RP^*B*^ and RP^*E*^, respectively (Fig. 7).

**Figure 7:**
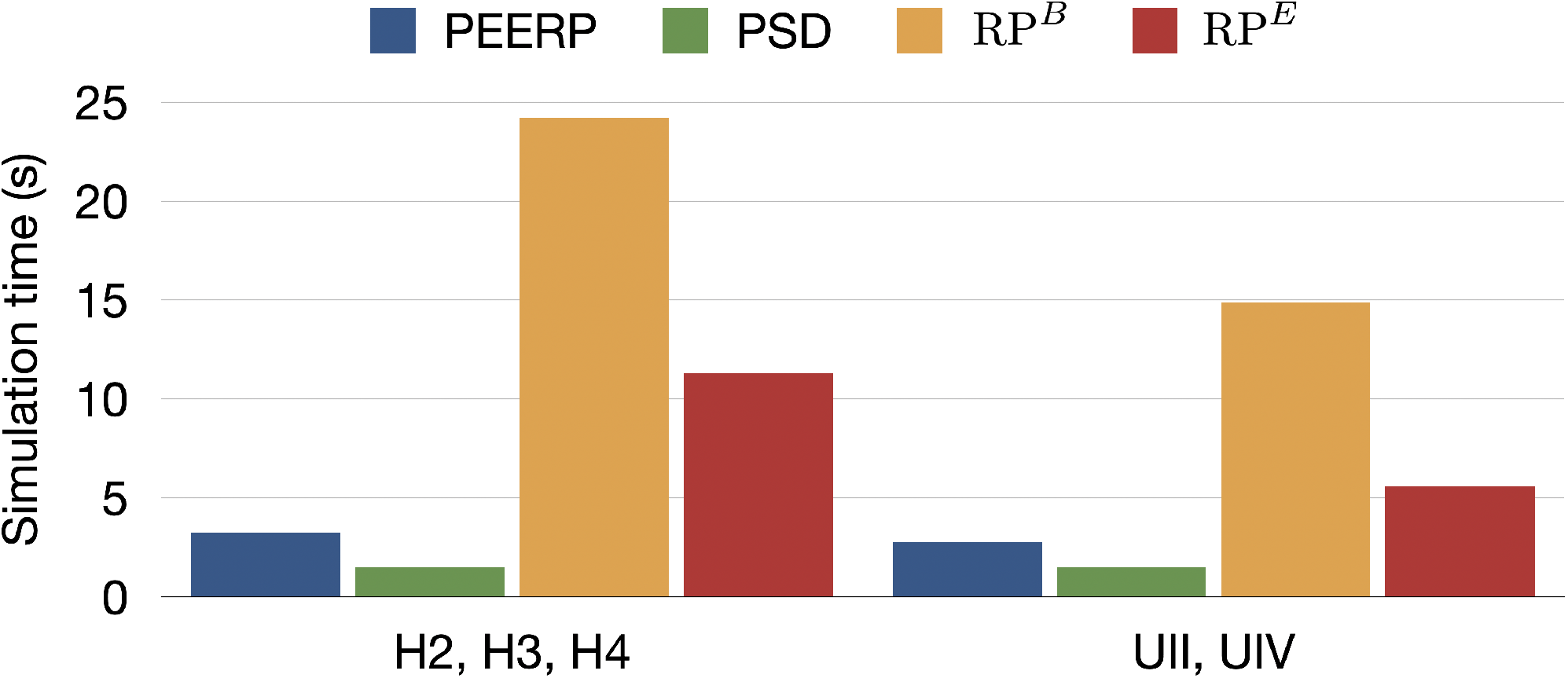
Simulation time needed to entirely perform each protocol from one stimulation point in ms.

Additionally, subsets of the initial pacing locations were extracted to evaluate the sensitivity of each protocol to a reduction of the number of pacing locations, i.e., an increase of the interpoint distance. The sensitivity was computed as the percentage of stable llPSs found when pacing from the initial pacing points with 1 cm inter-point distance. We considered increasing inter-point distances of 1.5, 2, 2.5 and 3 cm. The sensitivity analysis results are presented in Tab. 1. When applying the protocols only at pacing points located 3 cm away from each other, leading to a total of only 11 bi-atrial pacing sites, all the protocols could reproduce more than the 90% of the locations maintaining stable llPSs in the model UIV. The PEERP and the RP^*B*^ were the only methods to identify some of the stable llPSs sustaining areas with a greater inter-point distance when applied to all models.

**Table 1:** Sensitivity of each protocol in the different atrial models to increased inter-point distance.

| Atrial model | Protocol |  |  |  |
| --- | --- | --- | --- | --- |
|  | PEERP | PSD | RP<sup>B</sup> | RP<sup>E</sup> |
| H2 | 1.5 cm (83.3%) |  |  |  |
|  | 2 cm (83.3%) |  |  |  |
|  | 2.5 cm (83.3%) |  | 1.5 cm (100.0%) | 1.5 cm (100.0%) |
|  | 3 cm (33.3%) |  |  |  |
| H3 | 1.5 cm (50.0%) | 1.5 cm (42.9%) | 1.5 cm (62.5%) | 1.5 cm (16.7%) |
|  | 2 cm (37.5%) |  | 2 cm (25.0%) |  |
| H4 | 1.5 cm (22.2%) | 1.5 cm (81.8%) | 1.5 cm (57.1%) |  |
|  |  | 2 cm (36.4%) | 2 cm (7.1%) |  |
|  |  | 2.5 cm (36.4%) | 2.5 cm (7.1%) |  |
| UII | 1.5 cm (92.0%) | 1.5 cm (92.9%) | 1.5 cm (50.0%) | 1.5 cm (50.0%) |
|  | 2 cm (24.0%) | 2 cm (84.7%) |  |  |
|  | 2.5 cm (24.0%) | 2.5 cm (71.8%) |  |  |
|  |  | 3 cm (69.4%) |  |  |
| UIV | 1.5 cm (97.7%) | 1.5 cm (97.6%) | 1.5 cm (98.9%) | 1.5 cm (96.9%) |
|  | 2 cm (97.7%) | 2 cm (96.5%) | 2 cm (97.3%) | 2 cm (96.9%) |
|  | 2.5 cm (94.3%) | 2.5 cm (96.5%) | 2.5 cm (97.3%) | 2.5 cm (96.9%) |
|  | 3 cm (94.3%) | 3 cm (96.5%) | 3 cm (97.3%) | 3 cm (96.9%) |

### 3.2 Maintenance

All protocols showed a tendency for increasing number of segments in which stable llPSs were sustained in models with lower global CV or higher percentage of fibrotic remodelling (Fig. 8). PEERP was the protocol which sustained most stable llPSs in the models without structural remodelling. In the UII and UIV models, the PSD method was the one identifying more segments vulnerable to maintain stable llPSs, followed by PEERP. The RP protocols classified subsets of atrial segments found by the other methods as vulnerable to sustain stable llPSs in most of the cases.

**Figure 8:**
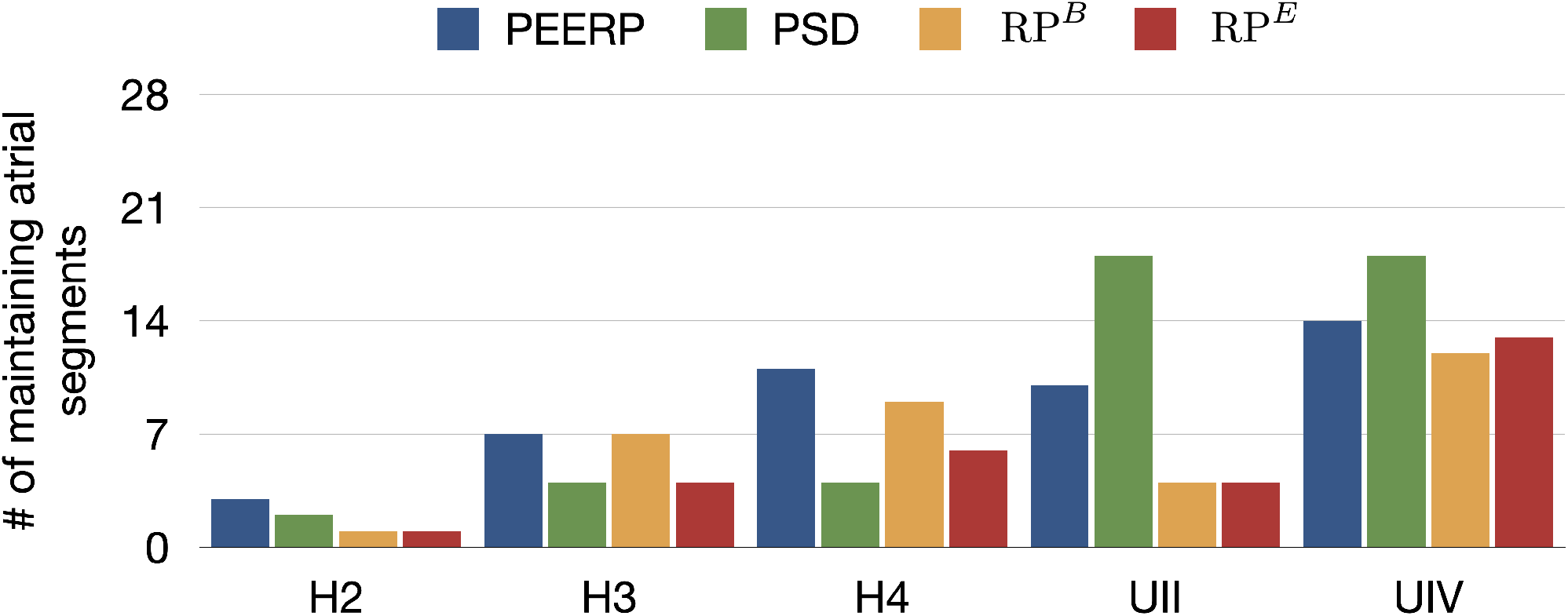
Number of atrial segments in which stable llPSs were sustained after applying the various protocols in the different models.

The PEERP method was the only one identifying the left atrial roof (segment 9) as vulnerable for maintenance of stable llPSs in all the models. All the pacing protocols (PEERP and RP) showed most of the stable llPSs in segments located in the left atrium. The inclusion of fibrotic remodelling (models UII and UIV) led to the maintenance of stable llPSs in the segments containing fibrosis. However, PSD classified as vulnerable to maintain many segments located in the right atrium too. Moreover, the PSD method yielded many stable llPSs sustaining in segments without fibrosis in the models UII and UIV. Independent of the choice of the inducibility protocol, most of the stable llPSs were maintained in the left posterior wall, roof, and septum (segments 5, 6, 9, 14, 16, 25, 27). The atrial segments in which stable llPSs were sustained after applying the different protocols can be found in the Supplementary Material.

The distance between the centers of the path travelled by stable llPSs and their corresponding inducing points was around 5-7 cm in most models using PEERP and RP^*B*^ (Fig. 9). Applying PSD and RP^*E*^ the distance was between 2.5 and 5 cm in the models without AF specific remodelling and between 5-7 cm in the others.

**Figure 9:**
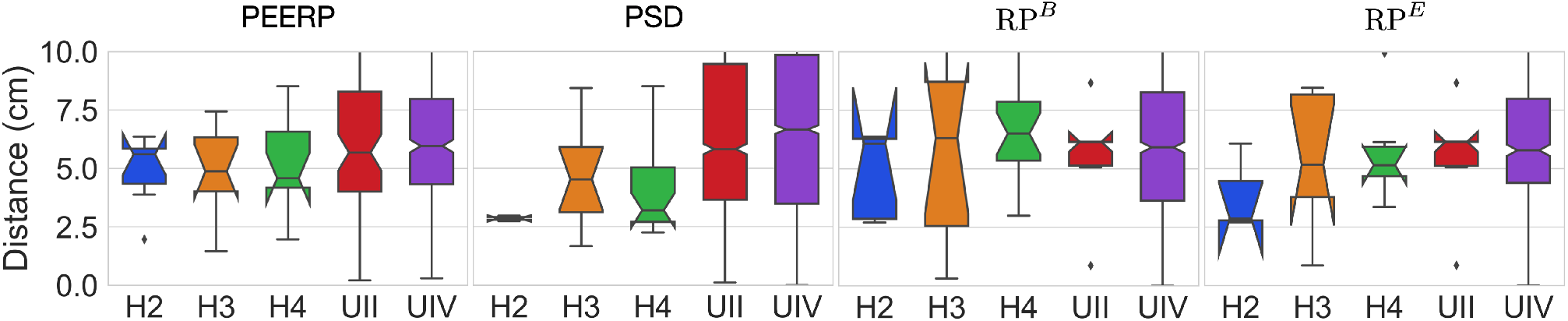
Geodesic distance between inducing points and centers of the path covered by stable llPSs.

## 4 Discussion

### 4.1 Inducibility

Each of the 4 protocols was applied at 227 locations for each of the 6 atrial models, resulting in 5,448 3D bi-atrial electrophysiological simulations. This is, to the best of our knowledge, one of the most extensive computational studies on the influence of pacing protocols on AF inducibility and maintenance. Regarding the vulnerability to induce arrhythmia, we demonstrated that the PEERP method provokes a bigger variety of different mechanisms compared to the other protocols. PEERP was able to induce diverse degrees of arrhythmia complexity, ranging from a flutter mechanism to chaotic multiple wave fronts. PEERP unveiled these complex mechanisms in most of the models which could potentially sustain them, even in the cases in which it was not the protocol with the highest number of inducing points. Furthermore, we showed that PEERP yields, overall, the highest number of segments from which arrhythmia could be induced (Fig. 5), meaning that this protocol is less dependent on the pacing location.

The results of the sensitivity analysis (Tab. 1), confirmed the ability of the methods PEERP to identify a good percentage of the total stable llPSs in all the models even with pacing locations placed at a greater inter-point distance between each other. These results are in line with [17], who could induce arrhythmia in 89% of patients with AF history with a train of stimuli close to the effective refractory period from a couple of pacing locations.

The application of the PEERP method showed that a similar number of beats (2-3) was needed to initiate arrhythmia in all the models used in this study. This result matches with the findings by [17], where three extra-stimuli were enough to initiate either AF or atrial flutter in patients both with and without history of AF. When fixing the maximum number of beats to 3, PEERP becomes a parameter-free method suitable to test inducibility in various electrophysiological models. Moreover, the smaller simulation time needed to perform the entire PEERP method from one stimulation point could give the possibility to test inducibility from more pacing locations to ensure the identification of all possible arrhythmic episodes which can arise in a given atrial model.

On the other hand, we showed a strong dependence of the induced episodes on the CI of the RP protocol (Fig. 6). Moreover, this parameter had to be tuned for each ionic model. Furthermore, we noticed that a single beat per CI is usually not enough to induce arrhythmia and then a second beat with the same CI is needed. More than 2 beats with the same CI rarely induced arrhythmia. This could confirm the clinical finding of [22], who found a protocol with decreasing CI (decremental pacing) to be more effective in inducing AF than a fast train of pulses with fixed CI (burst pacing).

Our study highlighted the importance of checking for arrhythmia initiation after every beat. Without this as a stopping criterion for the protocol, many arrhythmic episodes induced by previous stimuli were terminated by subsequent stimuli. Indeed, RP^*B*^ initiated arrhythmia before reaching the lowest CI and with a lower number of beats per CI than the predefined maximum *N* = 4 (Fig. 6). Finally, arrhythmic episodes were always more easily initiated by applying RP^*B*^ compared with RP^*E*^, however at a much higher computational cost. Our results are in line with clinical findings by [22], where inducibility differed depending on the method of induction, the number of induction attempts and the patient (in our case represented by different atrial models).

### 4.2 Maintenance

The areas identified as vulnerable to sustain reentries were different between the various protocols, confirming the importance of the choice of the induction protocol not only in initiation but also in maintenance of AF.

[17] were able to induce AF by applying stimuli close to the effective refractory period even in patients without previous AF history. In line with these findings, PEERP-induced stable llPSs were maintained in the models without fibrotic remodelling but slower global conduction velocity.

There was a strong correlation between maintenance of stable llPSs and presence of fibrotic tissue, mostly when applying pacing protocols (PEERP and RP). As shown in [4], the regions of the crista terminalis and pectinate muscles (located in segments 25 and 27) were prone to sustain AF in the right atrium due to their high degree of heterogeneity in fiber architecture and conduction velocity.

The PSD method labeled areas in the right atrium (segments 24 and 27) as highly vulnerable to sustain llPSs, which almost no other protocol found. This was because PSD is by definition setting PSs as initial conditions and could lead to PSs maintained close to the point in which they were initiated, mostly if not attracted by fibrotic tissue patches.

The arrhythmia check as stopping criteria played an important role not only in the initiation but also in the maintenance of stable llPSs, since it led to the identification of different segments as vulnerable to sustain reentry when comparing RP^*B*^ and RP^*E*^.

The average geodesic distance of 5-7 cm between endpoints of rotors and corresponding inducing points showed that, in most cases, stable llPSs sustain in areas not in the pacing location’s neighbourhood. Consequently, we needed to check arrhythmia initiation in the whole atria, not only in the proximity of the stimulation point.

The PEERP recognized the areas which we expected to be more vulnerable to sustain rotors, highly heterogeneous regions or containing fibrotic tissue. Moreover, the PEERP identified most of the segments classified as vulnerable to maintain stable llPSs by the other protocols too. This showed a high sensitivity and specificity of the PEERP method in discriminating between areas which are prone to sustain stable rotors.

### 4.3 Clinically Important Observations

We noticed stable llPSs dominating in atrial models including fibrotic tissue and perpetuating mostly in segments containing fibrosis. This confirmed the link between re-entrant drivers dynamics and the fibrotic tissue distribution [31], [44]. On the contrary, non-stable llPSs prevailed in atria without structural remodelling.

Moreover, we showed that not only fibrotic tissue distribution and conduction velocity, but also the induction protocol are influencing both initiation and progression of AF episodes, confirming what was clinically displayed by [22].

Different arrhythmic mechanisms were induced in our atrial models, with various degree of complexity. This could support that the rivalling theories of rotors [19] and multiple wavelet [32] were both right and can co-exist. Sometimes one mechanism dominates and sometimes the other.

### 4.4 Clinical Applicability of the PEERP Protocol

Rapid decremental pacing and burst pacing are considered as state-of-the-art protocols to induce arrhythmia in the atria [11], [22]. Moreover, suppression of inducibility with these protocol has been used as endpoint of ablation treatment by various groups [11], [12], [16]. However, the strong influence of the chosen protocol to initiate and maintain AF has been previously shown [22] and was confirmed mechanistically in our work. Furthermore, the lack of a consensus regarding which method to use to test inducibility makes studies hard or impossible to reproduce and to compare. Patient hearts are electrophysiologically diverse and human atrial myocardium is intrinsically heterogeneous, calling for a pacing protocol which is as much as possible independent from human-defined parameters (e.g. basic cycle length, coupling interval and decreasing step). A programmed atrial stimulation protocol [17] pacing close to the effective refractory period has shown positive predictive accuracy in inducing AF of 95% using only a few pacing locations and three atrial stimuli. We showed that an automatically adjustable pacing protocol stimulating at the end of the effective refractory period is able to initiate arrhythmia episodes with on average only 2-3 stimulations, in accordance with what was observed in [17]. [41] presented a method to assess atrial repolarization without provocative electrical stimuli using standard clinical catheters. This opens the possibility of applying our proposed PEERP method in clinical practice.

### 4.5 Limitations

Our atrial geometry did not include heterogeneous atrial wall thickness, which has been shown to influence the dynamics and stability of reentrant drivers [4]. However, we are confident that our results hold in presence of heterogeneity in atrial wall thickness, since the shown consistency of results with the PEERP on the variety of cases we investigated also translates to new variations. We carried out monodomain simulations which could have affected the dynamics of AF, even if the bidomain equations did not show significant difference in reproducing the wave propagation in thin-walled atrial tissue [35]. Due to the computational cost of this extensive study, we decided to limit the simulation time to 1.5 s after initiation of an arrhythmia. A meandering PS observed in our study could potentially stabilise later and affect the areas of maintenance.

## 5 Conclusion

Our study highlights the influence of different arrhythmia induction protocols for the assessment of both AF initiation and maintenance of AF. Our newly proposed PEERP protocol offers a reproducible, comprehensive and fast method to assess vulnerability. PEERP was able to provoke different degrees of arrhythmia complexity and unveil areas prone to maintain AF with a low number of stimuli, thus computationally inexpensive. The open source availability will facilitate adoption of the parameter-free PEERP method as a community standard. This work is a basis to increase comparability and reproducibility of in silico arrhythmia vulnerability studies and could prove feasible to be applied clinically as well.

## Data Availability

The methods used for this study are published open source as part of carputils and can are
explained in an example: www.opencarp.org/documentation/examples.

## Author Contributions

LA and AL conceived and designed the study. LA constructed the atrial models, implemented the protocols, ran the simulations, analyzed the data and drafted the manuscript. SS developed the method to subdivide the atria into segments and to compute the geodesic distance. All authors edited and approved the manuscript.

## Acknowledgments

The authors thank Jorge Patricio Arciniegas Sánchez and Laura Unger for their valuable suggestions and discussions. The authors thank Claudia Nagel for her precious feedback on the manuscript. This work was performed on the supercomputer ForHLR II funded by the Ministry of Science, Research and Arts of Baden-Württemberg and by the German Federal Ministry of Education and Research.

## Data Availability Statement

The methods used for this study are published open source as part of carputils and can are explained in an example: www.opencarp.org/documentation/examples.

