## Supplementary Data for "A Reproducible Protocol to Assess Arrhythmia Vulnerability in Silico: Pacing at the End of the Effective Refractory Period"

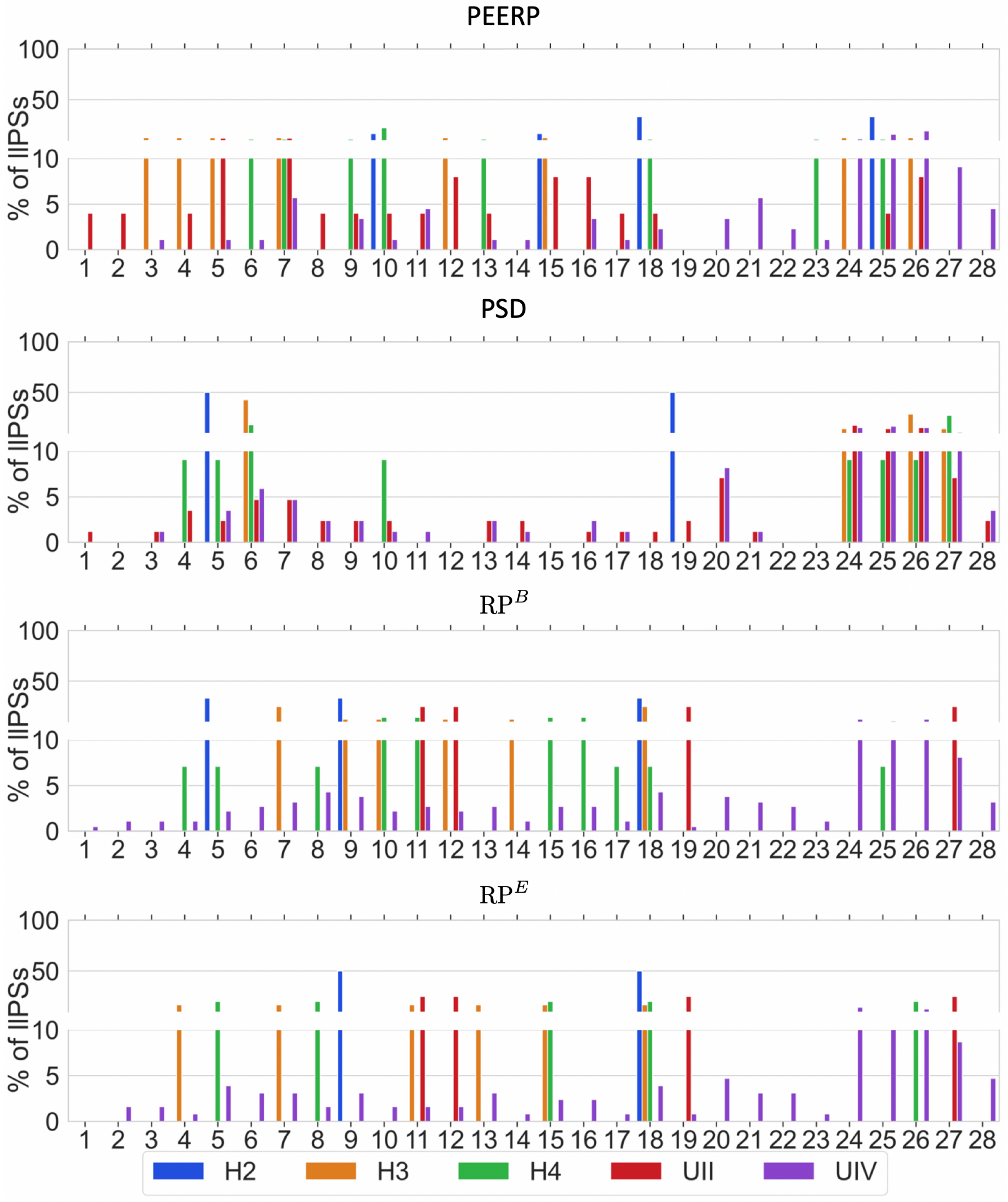


Figure 1: Atrial segments in which inducing points were identified applying the various protocols in the different models.

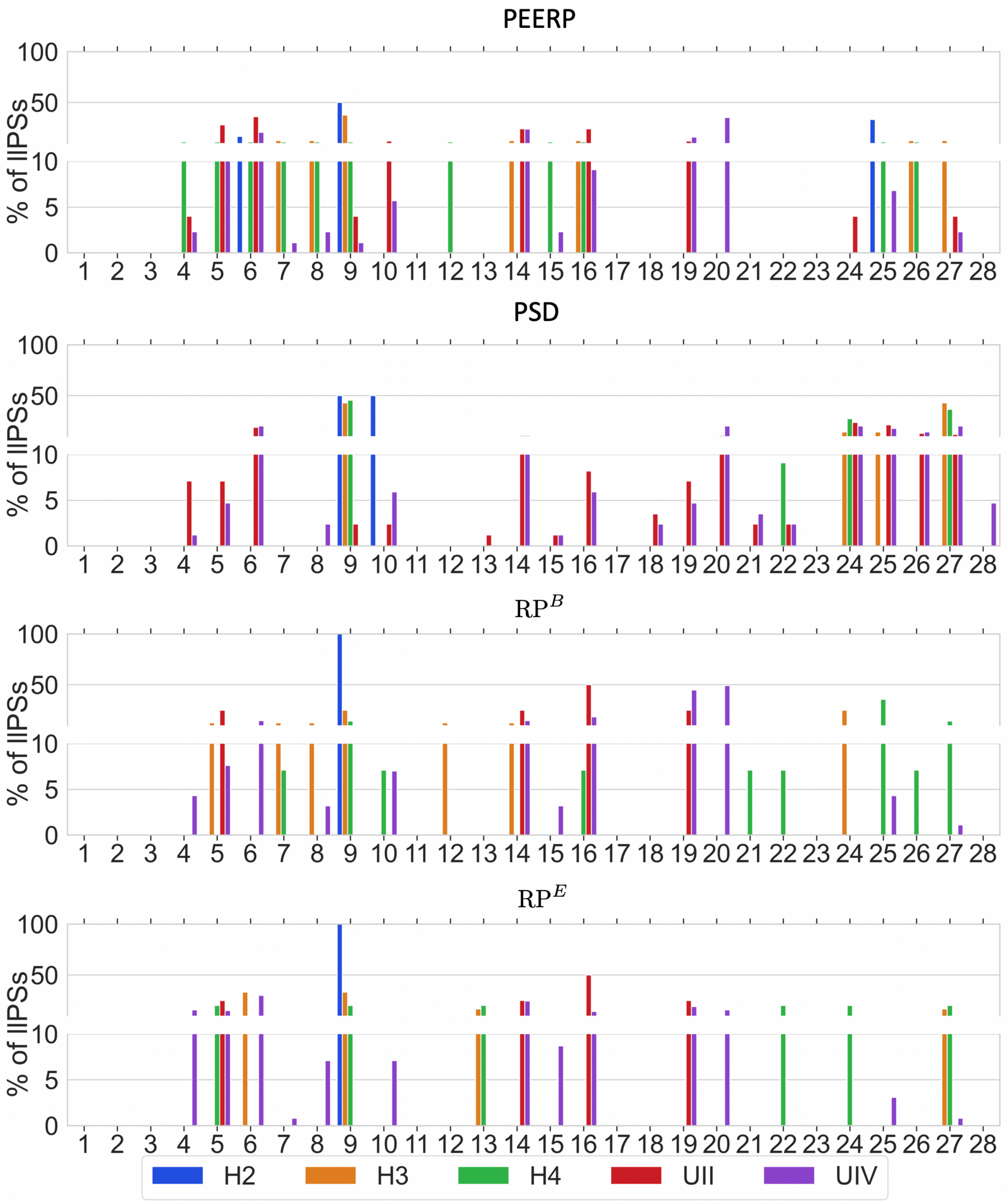


Figure 2: Atrial segments in which stable llPSs were maintained in the different models applying each protocol.
